# Clinical features of puff adder envenoming: case series of *Bitis arietans* snakebites in Kenya and a review of the literature

**DOI:** 10.1101/2024.05.31.24308288

**Authors:** Frank-Leonel Tianyi, Cecilia Ngari, Mark C. Wilkinson, Stanley Parkurito, Elizabeth Chebet, Evans Mumo, Anna Trelfa, Dennis Otundo, Edouard Crittenden, Geoffrey Maranga Kephah, Robert A Harrison, Ymkje Stienstra, Nicholas R Casewell, David G Lalloo, George O Oluoch

## Abstract

**Introduction:** The puff adder (*Bitis arietans*) is a medically important snake species found across much of Africa, yet there is a limited understanding of the clinical features and pathophysiology of envenoming after a puff adder bite.

**Methods:** We conducted a case-series study to describe the clinical features of patients with puff adder bites who were treated in two primary healthcare facilities in Kenya and complemented our case-series with a review of all published cases of puff adder envenoming that contained sufficient clinical details to highlight the major features.

**Results:** Between December 2020 and September 2021, 15 patients were admitted with a suspected puff adder bite (based on the patient’s description of the biting snake or confirmed in patients who brought the dead snake or a picture of the biting snake for identification) at the Chemolingot and Mwingi sub-county hospitals in central Kenya. Common local and systemic features on admission included pain (n=15, 100%), swelling (n=14, 93%), and haemorrhage (n=9, 60%). Coagulopathy (n=2, 13%) and shock (n=1, 8%) were less common. In addition, we conducted a literature review and identified 23 studies with detailed descriptions of the clinical features of puff adder envenoming from 37 patients. Local features were common and consistent across cases - swelling (100%, n=37) and pain (95%, n=35). Systemic features were less consistent, with 10 (27%) patients exhibiting hypotension on admission, 10 (27%) patients reporting a fever, and 13 (35%) developing anaemia. Some complications were common in patients with bites by captive snakes (amputations), compared to patients with bites by wild snakes (hypotension). Snake identification was easier and more accurate after bites by captive snakes, but for patients bitten in community settings, identification was challenging and often less objective.

**Conclusion:** We combined clinical cases and a literature review to describe the common and less common clinical features of puff adder envenoming. Further clinical research with serial laboratory assays of patients with definitively identified puff adder bites is crucial to further understand the pathophysiology of envenoming by this medially important snake species.

## Introduction

The puff adder (*Bitis arietans*) is one of the most medically important snakes in the world. It is a large terrestrial snake that hunts by ambush, has a broad geographic coverage (occurs naturally in 46 African and Middle Eastern countries), and a remarkable ability for camouflage[1]. It is commonly accidentally stepped upon by an unsuspecting victim in the savannah regions of sub-Saharan African countries, resulting in frequent bites to the lower limbs[2]. It is a popular attraction in zoos and among exotic pet traders. This expands the risk of a snakebite beyond the African continent, to Europe, North America, and Asia, where these snakes do not occur naturally.

Despite a high anecdotal burden of bites, there has been little empirical evidence detailing the epidemiological burden, or the clinical features of puff adder envenoming. Clinical studies have identified high proportions of puff adder bites among snakebite victims, for example, accounting for up to 75% of snakebites in Zimbabwe[3]. This study did not describe the clinical features of puff adder victims separately, precluding a deep understanding of the pathophysiology of puff adder envenoming. In 1975, Warrell et al[4] described the clinical details of 10 patients bitten by puff adders in the North of Nigeria, and almost 50 years later, this remains the most detailed study of puff adder envenoming in the academic literature. For an old-world viper with widespread geographic coverage, this is concerning because this single viewpoint on the features of puff adder envenoming may limit the recognition of puff adder envenoming in other regions, delay effective antivenom treatment, and increase the risks of adverse outcomes. This problem is accentuated by the heterogeneity in venom composition from puff adders within and between countries and regions[5]. The clinical, functional, and therapeutic consequences of other medically important snake species have been investigated and described[6–8], but not so for puff adders. This can be partly explained by the limited clinical research infrastructure in the primary health facilities that receive the most patients with puff adder bites, and the limited funding to support snakebite research[9, 10].

In this study, we aim to provide a comprehensive description of the clinical features of puff adder envenoming in patients treated in two primary healthcare facilities in Kenya and complement our case-series with a review of all published cases of puff adder envenoming that contained sufficient clinical details to highlight the major features of puff adder envenoming.

## Methods

Ethical approval for the case series portion of this study was obtained from the Kenyatta National Hospital – University of Nairobi and Ethics Research Committee, Nairobi, Kenya (P149/03/2020) and the Liverpool School of Tropical Medicine Research Ethics Committee (Research Protocol 18-058). Written informed consent was obtained from the participants prior to data collection. For participants less than 18 years of age, their assent was sought directly, and consent was obtained from their legal guardians. A witnessed consent process was used for patients who could not read or write.

### Case series

A clinical observational study was conducted in Baringo and Kitui Counties in Kenya, as part of the NIHR funded African Snakebite Research Group project. All patients who presented to the Chemolingot sub-county hospital (Baringo County) and the Mwingi sub-county hospital (Kitui County) with a suspected puff adder bite between XX December 2020 and XX September 2021 were reviewed by the study clinician who has expertise in managing puff adder envenoming. We included patients who presented within 24 hours of the bite and were ≥ 8 years old. Snake identification was by either visual inspection of the dead biting snake, a picture of the biting snake, or a detailed description of the snake to allow its identification by the treating team.

### Clinical evaluation

Case report forms were used to collect data on patient demographics, bite circumstances, bite characteristics, time of bite and time of hospital arrival, 20-minute whole blood clotting test (20WBCT) results, and details of antivenom treatment (other clinical indications, administration times, brand, dosage, adverse reactions).

### Statistical Analyses

Descriptive analysis of sociodemographic, bite circumstances and clinical data was carried out using proportions, means, and standard errors (SEs) for normally distributed variables and medians and interquartile ranges (IQRs) for nonparametric variables. Symptoms and signs were grouped into local and systemic envenoming and figures were created displaying the frequencies of each. Stata v16.1 was used for statistical analyses and a p-value <0.05 was used for statistical significance.

### Review

A single author (FLT) searched the PubMed database using the keywords (“puff adder”) OR (“*Bitis arietans*”), and the Global Index Medicus database using the keywords (tw:(puff adder)) OR (tw:(*Bitis arietans*)) on 15^th^ July 2023. All references, with no language restriction, were imported into EndNote version 20, duplicates were removed, and the references exported to Covidence systematic review software for screening and selectionn. Titles and abstracts were reviewed, and we included studies that involved human snakebite patients with no age restriction and described a clinical case or a series of cases of puff adder envenoming. Only clinical cases with information available on the snake species (confirmed or suspected), bite location, local and/or systemic symptoms and outcome were included. We excluded animal studies, in-vivo assays or observational studies in which symptoms and signs of envenoming were aggregated, preventing a detailed description of individual cases. Extracted data included last name of first author, year of publication, country where study was conducted, demographic data, salient clinical features, treatment, and outcome. Where available, detailed laboratory data were extracted and summarised as appropriate. We extended our search to include the reference lists of included articles, google scholar, and conference proceedings. The PRISMA-Scoping Review guidelines were used in the conduct and reporting of the review section of this paper.

## Results

### Clinical study

Between December 2020 and September 2021, 15 patients were admitted with a puff adder bite at the Chemolingot and Mwingi sub-county hospitals and enrolled into our study.

### Study population

The median age of study participants was 23.0 years (IQR: 16.0-35.0), and 53% were male. Patients had a median hospital delay of 8.1 hours (IQR: 5.3-11.3) after the snakebite. Herding was the most common occupation among patients (27%, n=4) and only two (13%) patients attended a traditional healer before hospital presentation. Most patients were walking outdoors when they were bitten (47%, n=7), though some were sleeping (33%, n=5), mostly outdoors, when they were bitten. The lower limbs were the most commonly bitten body part (73%, n=11).

### Biting snake identification

A combination of objective and subjective measures were used to identify the biting snake species. The biting snake was killed by the victim in three cases. One of the patients brought the dead snake to the hospital for identification, one patient took a picture of the dead snake and shared this with the healthcare workers, and the last patient provided a description of the snake and recognised the snake on a picture chart of medically important snakes in Kenya. Figure 1 shows pictures of the puff adders, and the bitten limb of one of the patients. The remaining 12 patients saw the biting snake, but neither brought a dead snake nor took a picture of the biting snake. The identification was made following a description of the biting snake, and a recognition of the snake on a picture chart of medically important snakes in Kenya. Despite being less objective, the latter identification method is worth considering because puff adders are very common in Kenya, they make a distinctive sound (puffing air from their body) before biting, and they are known locally as “*Kipsu*” in Baringo county. and *“Kimbuva”* in Kitui county.

**Figure 1.**
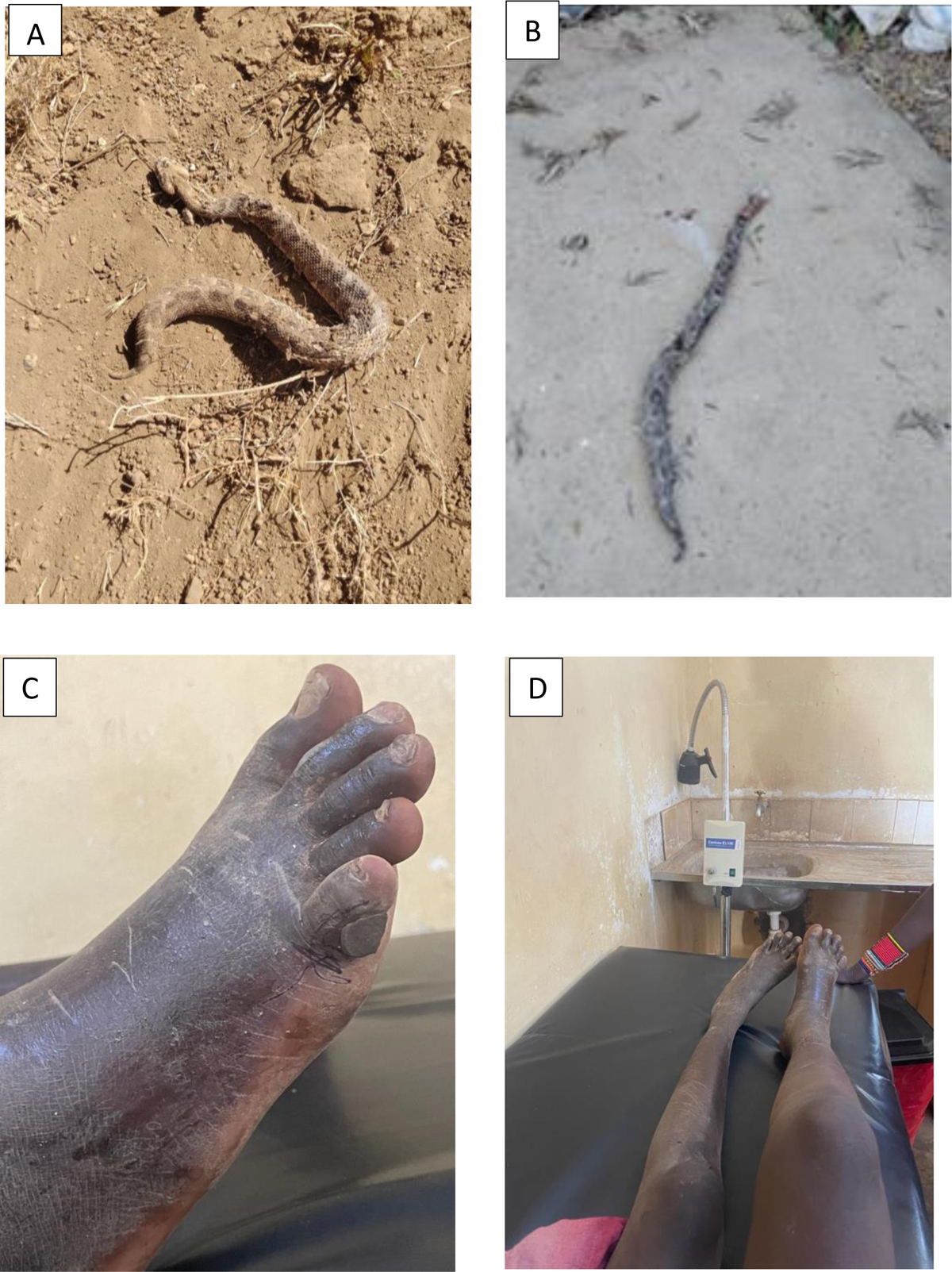
Biting snake and local effects of bite. A*. Picture of biting snake taken by snakebite victim and used for snake identification. B*. Picture of biting snake brought to the health facility for snake identification. C. Puff adder bite to the right small toe, and black stone applied as an early intervention (Patient bitten by snake in Figure B above). D. Progressive swelling of entire right leg after a puff adder bite to the right small toe (Patient bitten by snake in Figure B above) *Both snakes identified as *Bitis arietans* by LSTM herpetologist Edouard Crittenden and KSRIC herpetologist Geoffrey Kephah.

### Features of local and systemic envenoming

All 15 patients complained of pain on admission which was very severe in five (33%). There was an improvement in pain over time, with no persisting very severe pain, only one (7%) patient reporting severe pain, and seven patients reporting decreases in level of pain at 24 hours post admission. The most common local sign was swelling of the bitten part, present in 14 (93%) patients on admission, persisting through 12 hours in all, and still present in 12 (80%) at 24 hours post admission. Fang marks were visible in ten patients (67%) on admission, while six patients (40%) patients had bruises, and one patient had a blister. Bruises were more commonly detectable 6 hours after admission, with 10 (67%) patients having bruises at this time point.

Haemorrhage was the most common feature of systemic envenoming on admission, present in nine (60%) patients. The sites of haemorrhage were varied and included the bite site (n=8, 53%), gums (n=2, 13%) and venepuncture site (n=2, 13%), with some patients having more than one feature. At 6 hours, four (27%) patients still had signs of haemorrhage, with three (20%) bleeding from the bite site, and two (13%) bleeding from the gums. Coagulopathy, evidenced by an abnormal 20WBCT, was present in only two (13%) of patients on admission and one (7%) patient 24 hours later. Shock, defined as a systolic blood pressure less than 90 mmHg, was present in one (6%) patient on admission, and the low BP persisted at 6 hours, and resolved at 24 hours.

### Treatment and outcomes

All patients were treated with Inoserp Pan-Africa antivenom (500 LD_50_; a polyvalent anti-*Bitis* spp., *Dendroaspis* spp., *Echis* spp., and *Naja* spp. specific equine F(ab)’_2_ antivenom manufactured by Inosan Biopharma, Spain). Antivenom treatment indications were: rapid progressive swelling (RPS) extending to the fingers or toes in three patients, swelling involving more than two joints in two patients, swelling involving half of the limb in nine patients, and one patient having spontaneous bleeding [12]. Most patients had more than one feature of envenoming, but only one was recorded as the indication for antivenom (for example, the indication for two patients with an abnormal 20WBCT on admission were RPS and spontaneous bleeding). No patients required surgical intervention, and there were no fatal outcomes.

### Literature review

Our literature search returned 206 articles, of which eight were duplicates. After screening the titles and abstracts, 31 articles met our criteria for full text review, and 12 were excluded. We identified five further articles from our extended search of the reference lists of included articles, conference proceedings, and a cursory search of google scholar. We extracted and summarised data on 37 cases of puff adder bites, published in 23 studies between 1970 and 2023, nine (39%) studies were in sub-Saharan Africa[4, 13–20], six (26%) in North America[21–26], six (26%) in Europe[27–32], and two (9%) in Asia[33, 34]. The median age of all patients was 32.5 years (range 1 – 64), and 75% of patients were male. Bites were on the hands or fingers in 77% of cases, and this was most common among snake handlers in non-African countries. Figure 2 shows the PRISMA flow chart on studies in this review, while Table 2 presents a summary of sociodemographic and clinical features from the included articles.

**Table 1.** Characteristic of puff adder patients in Kenyan clinical case series.

| **Patient characteristics** | **Total (N=15)** |
| --- | --- |
| Age, median (IQR) | 23.0 (16.0-35.0) |
| Gender |  |
| Male | 8 (53%) |
| Female | 7 (47%) |
| Occupation |  |
| Herder | 4 (27%) |
| Students | 1 (7%) |
| Housewife | 2 (13%) |
| Unemployed | 3 (20%) |
| Unknown | 5 (33%) |
| Activity when bitten |  |
| Outdoors/Walking | 7 (47%) |
| Toilet visit | 1 (7%) |
| Sleeping | 5 (33%) |
| Indoors | 2 (13%) |
| Bite location |  |
| Foot | 6 (40%) |
| Ankle/knee | 5 (33%) |
| Hand/Forearm | 2 (13%) |
| Arm/Shoulder | 1 (7%) |
| Face | 1 (7%) |
| Visited a traditional healer |  |
| Yes | 2 (13%) |

**Table 2.** Summary of sociodemographic and clinical features of published cases of puff adder envenoming.

| No | Author | Year | Location | Number of cases | Age (yrs)/ Median age(range) | Gender | Captive/ Wild snake | Bite sites | Clinical signs and symptoms | Antivenom name (%) | Surgical care/ % | Outcome |
| --- | --- | --- | --- | --- | --- | --- | --- | --- | --- | --- | --- | --- |
| 1 | Takahashi et al[26] | 1970 | USA | 1 | 30 | Male | Captive | Right index finger | Fang marks<br>Local bleeding<br>Severe pain<br>Progressive swelling<br>Swollen tender axillary lymph nodes<br>Fever | SAIMR polyvalent (100%) | Nil | Full recovery |
| 2 | Sezi et al[19] | 1972 | Uganda | 2 | 13<br>50 | Male<br>Male | Wild | Right foot (50%)<br>Left hand (50%) | Fang marks (50%)<br>Severe swelling (100%)<br>Hypotension (100%)<br>Anaemia (100%)<br>Local bleeding (100%)<br>Distal haematoma (50%)<br>Fever (50%) | Polyvalent anti-snake serum (100%) | Surgical debridement and skin grafting (50%) | Full recovery (100%) |
| 3 | Warrell et al[4] | 1975 | Nigeria | 10 | 24.5 (9 – 55) | NR | Wild | Calf (40%)<br>Ankle (20%)<br>Heel (10%)<br>Toe (10%)<br>Hand (10%)<br>Thumb (10%) | Fang marks (40%)<br>Local bleeding<br>Bleeding gums (10%)<br>Epistaxis (20%)<br>Hypotension (20%)<br>Severe pain (100%)<br>Progressive swelling (70%)<br>Blisters (50%)<br>Fever (50%)<br>Vomiting (30%)<br>Tender lymph nodes (10%) | SAIMR polyvalent (70%) | NR | Deaths (20%) |
|  |  |  |  |  |  |  |  |  | Jaundice (20%)<br>Anaemia (30%) |  |  |  |
| 4 | Reid[28] | 1978 | UK | 5 |  | NR | Captive | Left middle and index fingers (20%)<br>Left hand (20%)<br>NR (60%) | Severe pain<br>Progressive swelling (20%)<br>Systemic poisoning (40%)<br>Mild poisoning (20%) | Antivenom name not given (80%) | NR | Necrosis (60%) |
| 5 | Sigel et al[29] | 1980 | Germany | 1 | 29 | Male | Captive | Right thumb | Progressive swelling<br>Local bleeding<br>Blisters<br>Signs of compartment syndrome | Polyvalent antivenom | Fasciotomy of the forearm | Full recovery |
| 6 | Schweitzer et al[18] | 1981 | South Africa | 1 | 16 | Female | Wild | Left index finger<br>Right hand | Fang marks<br>Local bleeding<br>Severe pain<br>Progressive swelling<br>Clinical diagnosis of compartment syndrome in left forearm<br>Bilateral carpal tunnel syndrome | SAIMR polyvalent (100%) | Bilateral carpal tunnel release<br>Fasciotomy of the forearm | Full recovery |
| 7 | Brossy et al[14] | 1982 | South Africa | 1 | NR | Male | NR | Left index finger | Fang marks<br>Progressive swelling | Polyvalent antivenom | Nil | Full recovery |
| 8 | Hamby et al[24] | 1983 | USA | 1 | 29 | Male | Captive | Right hand (thenar region) | Fang marks<br>Severe pain<br>Progressive swelling | Antivenom name not given | Nil | Full recovery |
| 9 | Theakston et al[31] | 1985 | UK | 1 | 21 | Male | Captive | Left fifth finger | Severe pain<br>Progressive swelling<br>Tender swollen axillary lymph nodes<br>Fever | No antivenom | Nil | Full recovery |
| 10 | Pugh et al[17] | 1987 | Nigeria | 2 | 35 (20 – 50) | Female | Wild | Toe (50%)<br>Foot (50%) | Severe pain<br>Progressive swelling (50%) | SAIMR polyvalent (50%) | Nil | Full recovery |
| 11 | Bey et al[21] | 1997 | USA | 1 | 35 | Male | Captive | Right index finger | Fang marks<br>Local bleeding<br>Severe pain<br>Progressive swelling<br>Ecchymosis | Behringwerke polyvalent (100%) | Debridement | Cutaneous scarring<br>Complete loss fingernail |
| 12 | Lavonas et al[25] | 2002 | USA | 1 | 37 | Female | Captive | Right ring finger | Haemorrhagic vesicles<br>Hypotension<br>Ecchymosis<br>Severe pain<br>Progressive swelling<br>Local bleeding<br>Necrotic finger<br>Fever<br>Anaemia | SAIMR polyvalent (100%) | Digit amputation | Digit amputation |
| 13 | Le Dantec et al[16] | 2004 | Senegal | 1 | 24 | Male | Wild | Right calf | Severe pain<br>Progressive swelling<br>Local bleeding<br>Hypotension<br>Bleeding from phlebotomy sites | Fav-Afrique polyvalent | Debridement | Full recovery |
|  |  |  |  |  |  |  |  |  | Blisters<br>Bullae<br>Necrosis |  |  |  |
| 14 | Strubel et al[30] | 2008 | Germany | 1 | 20 | Male | Captive | Right index finger | Severe pain<br>Progressive swelling<br>Clinical diagnosis of compartment syndrome in the right forearm<br>Signs of impaired coagulation<br>Necrosis | Antivenom name not given | Debridement<br>Fasciotomy<br>Digit amputation | Loss of finger |
| 15 | Disser et al[22] | 2010 | USA | 1 | 43 | Male | Captive | Right index finger | Fang marks<br>Severe pain<br>Progressive swelling<br>Finger necrosis | SAIMR polyvalent | Digit amputation | Loss of finger |
| 16 | Rainer et al[27] | 2010 | Austria | 1 | 26 | Male | Captive | Right hand | Fang marks (4)<br>Severe pain<br>Progressive swelling<br>Erythema<br>Hypotension | SAIMR polyvalent | Nil | Full recovery |
| 17 | Dymarsky et al[23] | 2015 | USA | 1 | 50 | Female | Captive | Neck (left submandibular region) | Severe pain<br>Progressive swelling<br>Anaemia | SAIMR polyvalent | NR | Full recovery |
| 18 | Firth et al[15] | 2016 | South Africa | 1 | 1 | Male | NR | Left upper limb | Severe pain<br>Progressive swelling<br>Clinical diagnosis of compartment syndrome<br>Coagulopathy + Anaemia | SAIMR polyvalent | Fasciotomy of the arm and forearm | Death |
|  |  |  |  |  |  |  |  |  |  |  | Debride-<br>ment |  |
| 19 | Benjamin<br>et al[13] | 2020 | Guinea | 1 | 64 | Male | Captive | Left<br>index<br>finger | Severe pain<br>Progressive swelling<br>Local bleeding<br>Blisters<br>Coagulopathy | Inoserp<br>Pan-Africa<br>polyvalent<br>antivenom<br>2 vials<br>Pt declined<br>further<br>vials | Nil | Full<br>recovery |
| 20 | Engelbrecht[32] | 2021 | Ireland | 1 | 22 | Male | Captive | Right hand<br>(first<br>web<br>space) | Severe pain<br>Progressive swelling | SAIMR<br>polyvalent | Nil | Full<br>recovery |
| 21 | Wakasugi<br>et al[34] | 2021 | Japan | 1 | 23 | Male | Captive | Left<br>middle<br>finger | Severe pain<br>Progressive swelling<br>Low blood pressure<br>Subcutaneous<br>haemorrhage in left arm<br>and back | Nil | Nil | Mild<br>contractur<br>es of the<br>middle<br>and ring<br>fingers |
| 22 | Husain et<br>al[33] | 2023 | Malaysia | 1 | Mid-30s | Male | Captive | Dorsum<br>of both<br>hands | Fang marks<br>Severe pain<br>Progressive swelling<br>Hypotension<br>Coagulopathy<br>Anaemia<br>Distal subcutaneous<br>haemorrhage | Nil | Nil | Death |
| 23 | Smit et<br>al[20] | 2023 | South<br>Africa | 1 | 27 | Male | NR | Left<br>wrist | Fang marks<br>Severe pain<br>Progressive swelling | Polyvalent<br>antivenom | Nil | Full<br>recovery |
|  |  |  |  |  |  |  |  |  | Clinical diagnosis of<br>compartment syndrome |  |  |  |

**Figure 2:**
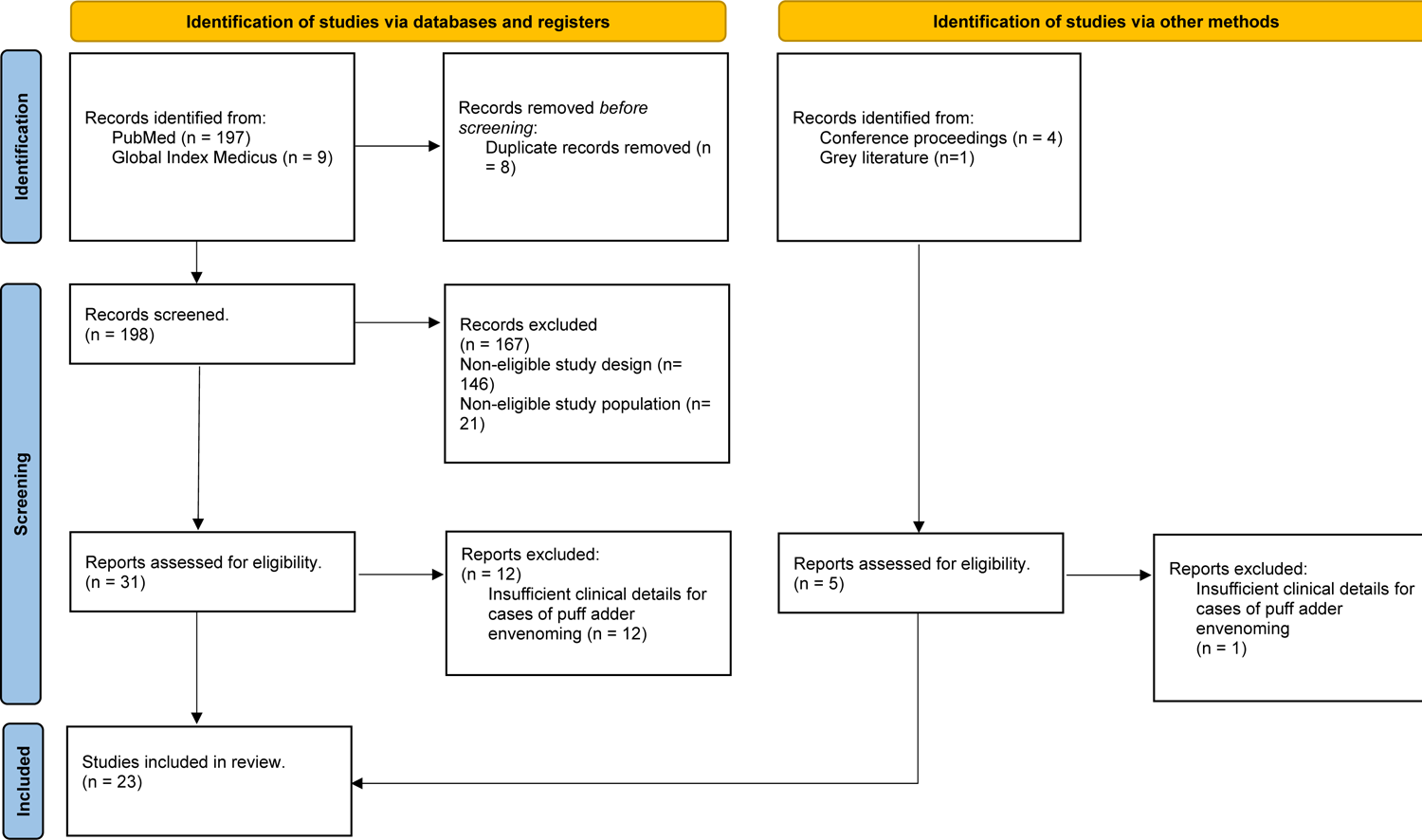
PRISMA flow chart of studies assessed for this review

### Biting snake identification

The methods of snake identification reported in the literature differed for patients between and within studies. The most accurate and direct methods were by expert identification of exotic snakes that bit their owner (n=12, 32%)[21–24, 26–28, 30–34], patients that brought the dead snake to the hospital (n=9, 24%)[4, 18] and snakes being handled in professional settings (n=7, 18%)[13, 19, 25, 28]. *Bitis arietans* venom was detected in serum samples of patients using ELISA techniques for four patients (11%)[4, 17]. For two patients (5%)[16, 19], the biting snake was identified based on the patient’s description, and for three patients (8%) the method of identification was not reported[14, 15, 20]. Overall, 32 (86%) patients had an objectively confirmed puff adder bite, a majority of which were bites by captive snakes, while five (14%) cases were subjectively identified, all of which occurred on the African continent, most likely from bites by wild snakes

### Clinical features of puff adder envenoming reported in the literature

Severe swelling and pain were the most commonly reported local features of puff adder envenoming in the literature, respectively reported by 37 (100%) [4, 13–34] and 35 (95%)[4, 13, 15–18, 20–28, 30–34] patients. Common local features included fang marks and local bleeding, which were reported respectively by 22 (69%) [4, 14, 18–22, 24, 26, 27, 33] and 23 (62%) [4, 13, 16, 18, 19, 21, 25, 26, 29] cases. Blisters, necrosis, and ecchymosis were uncommon local features, occurring in eight (22%) [4, 13, 16, 29], six (16%) [4, 13, 16, 29] and four (11%)[21, 25] cases respectively. Three patients [4, 26, 31] developed a tender swelling of the lymph nodes proximal to the bitten body part. One developed a tender swelling in the right axillary and right inguinal lymph nodes after a bite to the right hand[4]. In six (16%) [15, 16, 18, 20, 29, 30] cases, investigators suspected a compartment syndrome and four [15, 18, 29, 30] of these underwent a fasciotomy.

Systemic features were less consistent across the reported studies. Ten (27%) patients across seven studies[4, 16, 19, 25, 27, 33, 34] had hypotension on admission, with blood pressures as low as 60/30 mmHg[19]. The hypotension resolved after intravenous (IV) fluids combined with antivenom treatment, but it persisted in the two patients who received only IV fluids without antivenom, with fatal outcomes[4, 33]. Ten (27%) patients across six studies[4, 15, 17, 19, 25, 29] reported a fever, with temperature measurements as high as 40.5 °C, and some patients remaining febrile up to 8 days after the bite. Eight of these patients[4, 17, 19, 29] were febrile on admission, while two patients[15, 25] developed a fever later during their hospitalisation. Four of these patients[4, 15, 25] developed local necrosis at the bite site, but there was not enough detail in the articles to confirm or exclude an infected bite wound as the cause of the fever. Thirteen (35%) patients across nine studies[4, 16, 17, 19, 23, 25, 30, 33, 34] had clinical or laboratory evidence of anaemia, and some had haemoglobin concentrations as low as 5.1 g/dl[19] on arrival. In some cases, the anaemic patients had a normal 20WBCT[4]. All 13 patients had local bleeding from the bite site, and in one patient, the bleeding persisted for three days after the bite. This patient had been admitted and treated with antivenom two days after the bite, and the local bleeding stopped within 24 hours of antivenom treatment[19]. Two patients, who received appropriate antivenom within six hours of the puff adder bite, developed haemorrhagic vesicles close to the bite site, and had an acute drop in haemoglobin concentration (14.3 -> 7.1g/dl[25], and 10.0 -> 7.4g/dl[4]) within eight and 35 hours of the puff adder bite respectively. Two patients had a haemoglobin drop with no external evidence of bleeding (13.4 -> 9.5g/dl[23] and 14.2 -> 9.7g/dl[34]) within 48 hrs of the bite. Some patients developed spontaneous haemorrhage at sites other than the bite wound. The spontaneous haemorrhage was sometimes obvious; for example bleeding gums and epistaxis after a bite to the calf[4]) but could be concealed (a hematoma in the jaw after a bite to the foot[19], and two cases of subcutaneous haemorrhage in the back following bites to the hand[33, 34]). One case of spontaneous haemorrhage into the aortic adventitia was detected after an autopsy of a fatal puff adder bite[4]. Five (14%) patients [13, 15, 16, 30, 33] had signs of coagulopathy, an abnormal 20WBCT in four cases[13, 15, 30, 33], and prolonged bleeding from a phlebotomy site in one case[16]. Seven (19%) patients[4, 16, 17, 19, 33] were reported to have received a transfusion of blood products (whole blood or packed red blood cells) due to acute anaemia after a puff adder bite.

### Treatment and outcomes reported in the literature

Antivenom use varied considerably within and between studies. Five (14%) patients across three studies did not receive antivenom, despite other patients in these same studies receiving antivenom[4, 17, 28]. Four of these patients were reported to have no indication for antivenom use, and for one patient there was no antivenom in the health facility. In three other cases, no antivenom use was reported at all[31, 33, 34]: in one case the patient only had mild local signs of envenoming[31], in the two remaining cases, the clinicians were unable to access antivenom[33, 34]. Overall, eight (28%) patients did not receive antivenom, with two of them having a fatal outcome[4, 17, 28, 31, 33, 34]. South African Institute for Medical Research (SAIMR) antivenom (a polyvalent anti *Bitis* spp., *Dendroaspis* spp., *Hemachatus* spp. and *Naja* spp. specific fragmented equine immunoglobulin [F(ab)’_2_] based antivenom manufactured by South African Vaccine Producers [SAVP], South Africa) was administered to 11 (30%) patients[4, 17, 18, 22, 23, 25, 26, 28, 32], with doses ranging from 20 – 80 ml. Two patients had an immediate adverse reaction, characterised by wheezing and hypotension[17, 25]. The occurrence of late serum reactions was not reported. Schlangengift-Immunserum antivenom (a discontinued polyvalent anti *Bitis* spp., *Echis* spp., and *Naja* spp. polyvalent equine antivenom, previously manufactured by Behringwerke, Germany) was administered to eight patients[4, 19, 21]. Inoserp Pan-Africa (500 LD50)[13], Fav-Afrique (a polyvalent anti *Bitis* spp., *Dendroaspis* spp., *Echis* spp., and *Naja* spp. specific equine F(ab)’_2_ antivenom formerly manufactured by Aventis Pasteur, France)[16] and FitzSimons’s antivenomous serums (a discontinued polyvalent anti *Bitis* spp., *Hemachatus* spp., and *Naja* spp. equine antivenom)[4] were each used to treat one patient. The name of the antivenom was not recorded for seven patients, but the authors reported using a polyvalent antivenom[20, 24, 28–30]. No data was reported on adverse reactions for the other antivenoms.

Surgical interventions were performed on 15 (41%) patients[4, 15, 16, 18, 19, 21, 22, 25, 28–30]. Eleven (30%) patients developed necrosis at the bite site, occurring between 24 hours and 1 week after the bite[4, 19, 21, 22, 25, 28, 30]. A surgical debridement was performed on nine (11%) patients[4, 15, 16, 19, 21, 28, 30], and four (11%) patients had a digit amputation[22, 25, 28, 30], with the amputation being preceded by a debridement in three cases[22, 25, 30]. A fasciotomy was performed on four patients for a suspected compartment syndrome[15, 18, 29, 30]. No loss of function was described in surviving patients, and only one patient required skin grafting[19].

There were four (11%) fatal cases in our review[4, 15, 33]. Two of these patients did not receive antivenom and died from persistent hypotension and severe anaemia, resistant to IV fluids and the transfusion of blood products[4, 33]. The remaining two were treated with SAIMR polyvalent antivenom; one patient died from repeated lower respiratory tract infections and overwhelming sepsis[15], while the second patient developed post operative complications (severe electrolyte imbalance and ventricular fibrillation) after a delayed amputation of his right leg for gangrene[4].

### Captive vs wild puff adder envenoming

We compared the major clinical features of envenoming following bites by wild (15 patients) or captive (19 patients) snakes to explore any patterns that can influence management and outcomes. Captive bites were more common on the digits (68%)[21, 22, 25, 26, 28–31, 34], compared to wild bites in which the bite sites were more varied. Compared to patients with captive bites, patients with wild bites had a higher documented occurrence of hypotension (21%[25, 27, 33, 34] vs 40%[4, 16, 19]), fever (11%[25, 29] vs 47%[4, 17, 19]), and anaemia (26%[23, 25, 30, 33, 34] vs 47%[4, 16, 17, 19]). The occurrence of bite site necrosis (32%[22, 25, 28, 30] vs 33%[4, 16, 19]) was similar in both groups, while patients with captive bites had a higher occurrence of amputations (21%[22, 25, 28, 30] vs 7%[4]). These differences were not statistically significant (p=0.35), and Figure 3 below compares the absolute number of patients with each of the above features or complications of puff adder bites.

**Figure 3.**
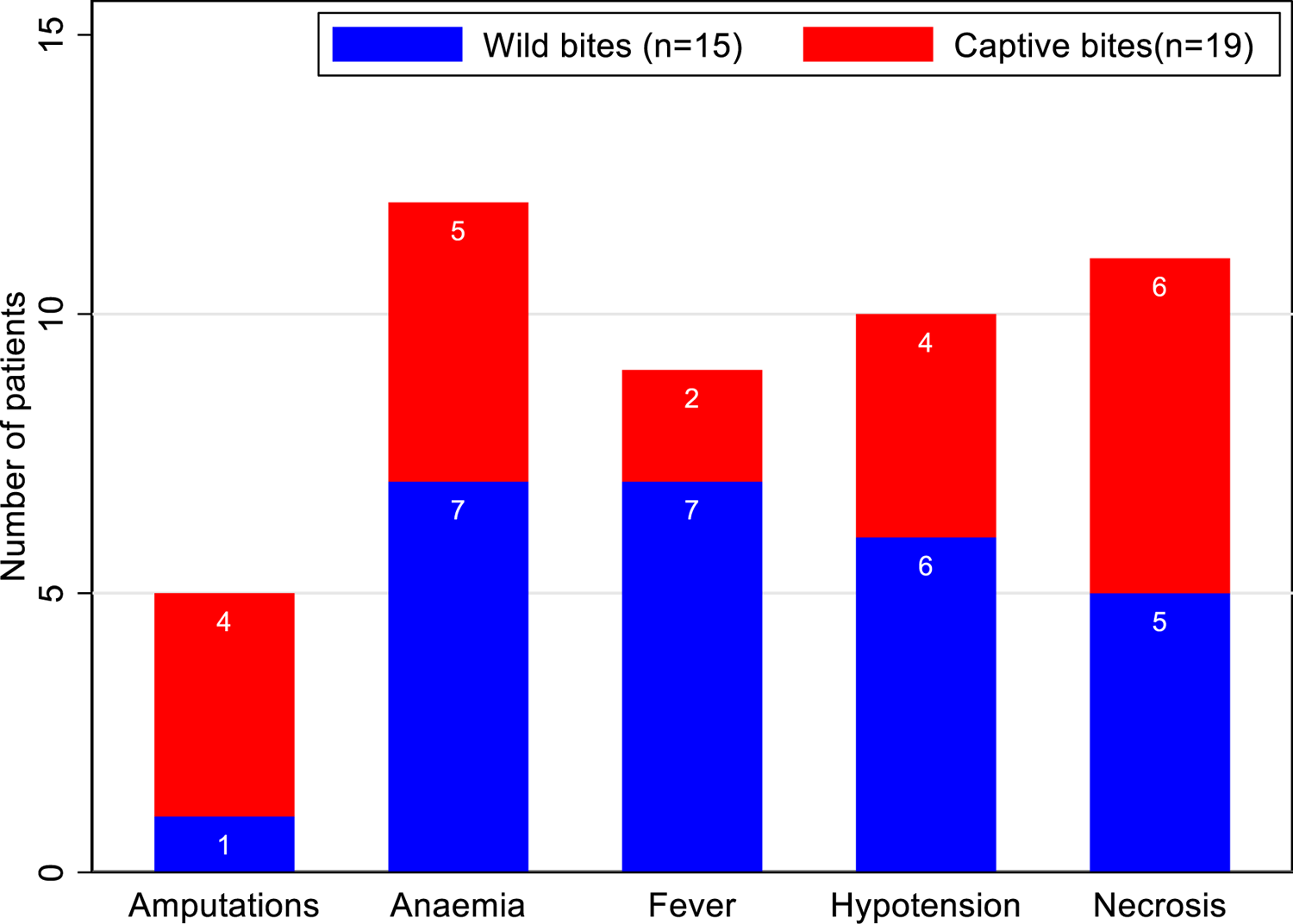
Clinical features and complications of puff adder bites in patients with wild and captive bites. *Some patients had more than one feature or complication. The origin of the snake was not reported in three cases

### Laboratory features of puff adder envenoming reported in the literature

Serial laboratory measures were only reported in three patients to allow an assessment of evolution of laboratory features of envenoming, and these are presented in table 3. There was a haemoglobin drop of 2.3g/dl[21], 4.5g/dl[34] and 4.6g/dl[33] in all three patients. All patients had at least one episode of thrombocytopenia, with levels as low as 34,000 cells per microliter in one patient[33]. Only one patient had a marked leucocytosis, with white blood cell count increasing from 6400 to 17500 cells per microliter[33]. Coagulation assays were mildly abnormal. The fibrinogen concentration remained normal in all three patients, while the prothrombin time/INR and APTT were normal in one patient[21], mildly raised in a second (max INR 1.38)[34], and more raised in the third (max INR 2.93)[33]. Serum electrolyte results were available for only one patient[33], and they had a severe hypokalaemia (2.5 millimoles per litre) on admission, and an acute kidney injury with a progressive increase in serum creatinine levels from 68.8 to 261.3 micromoles per litre. Serial creatine kinase levels were reported for two patients[33, 34] and these increased progressively over time, ranging between 90 and 330 units per litre.

**Table 3.**
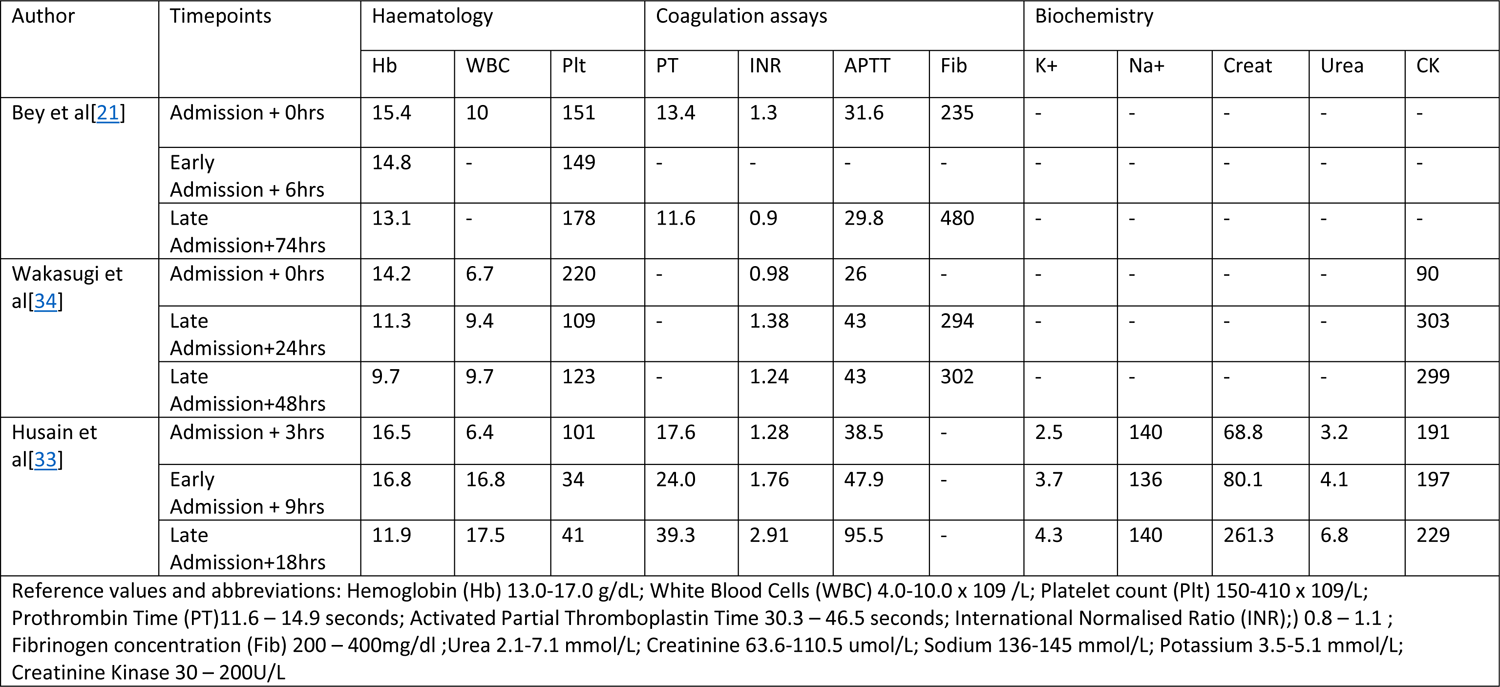
Serial laboratory assays of published cases of puff adder envenoming.

| Author | Timepoints | Haematology |  |  | Coagulation assays |  |  |  | Biochemistry |  |  |  |  |
| --- | --- | --- | --- | --- | --- | --- | --- | --- | --- | --- | --- | --- | --- |
|  |  | Hb | WBC | Plt | PT | INR | APTT | Fib | K+ | Na+ | Creat | Urea | CK |
| Bey et al[21] | Admission + 0hrs | 15.4 | 10 | 151 | 13.4 | 1.3 | 31.6 | 235 | - | - | - | - | - |
|  | Early Admission + 6hrs | 14.8 | - | 149 | - | - | - | - | - | - | - | - | - |
|  | Late Admission+74hrs | 13.1 | - | 178 | 11.6 | 0.9 | 29.8 | 480 | - | - | - | - | - |
| Wakasugi et al[34] | Admission + 0hrs | 14.2 | 6.7 | 220 | - | 0.98 | 26 | - | - | - | - | - | 90 |
|  | Late Admission+24hrs | 11.3 | 9.4 | 109 | - | 1.38 | 43 | 294 | - | - | - | - | 303 |
|  | Late Admission+48hrs | 9.7 | 9.7 | 123 | - | 1.24 | 43 | 302 | - | - | - | - | 299 |
| Husain et al[33] | Admission + 3hrs | 16.5 | 6.4 | 101 | 17.6 | 1.28 | 38.5 | - | 2.5 | 140 | 68.8 | 3.2 | 191 |
|  | Early Admission + 9hrs | 16.8 | 16.8 | 34 | 24.0 | 1.76 | 47.9 | - | 3.7 | 136 | 80.1 | 4.1 | 197 |
|  | Late Admission+18hrs | 11.9 | 17.5 | 41 | 39.3 | 2.91 | 95.5 | - | 4.3 | 140 | 261.3 | 6.8 | 229 |
| Reference values and abbreviations: Hemoglobin (Hb) 13.0-17.0 g/dL; White Blood Cells (WBC) 4.0-10.0 x 10<sup>9</sup> /L; Platelet count (Plt) 150-410 x 10<sup>9</sup>/L; Prothrombin Time (PT)11.6 – 14.9 seconds; Activated Partial Thromboplastin Time 30.3 – 46.5 seconds; International Normalised Ratio (INR);) 0.8 – 1.1 ; Fibrinogen concentration (Fib) 200 – 400mg/dl ;Urea 2.1-7.1 mmol/L; Creatinine 63.6-110.5 umol/L; Sodium 136-145 mmol/L; Potassium 3.5-5.1 mmol/L; Creatinine Kinase 30 – 200U/L |  |  |  |  |  |  |  |  |  |  |  |  |  |

## Discussion

Despite being a medically important snake with a wide geographical distribution, puff adder envenoming has been rarely described in the published literature. Our review found only 37 cases that have been described since 1970 with sufficient detail to understand the local and systemic features of puff adder envenoming, with only three of these cases being reported with serial laboratory assays. We added this review of published cases to the clinical reports of 15 cases of puff adder envenoming from two hospitals in Kenya reported in this study.

### Geographic specificities

Puff adders are native to the African continent and the Middle East. In our literature review, 54% (20/37) of previous cases of envenoming occurred in sub-Saharan Africa. The profile of patients presenting with a puff adder bite on the continent was heterogenous, including farmers, school children, and herpetologists. From our case series, the most common formal occupation was herding, and a significant proportion of patients had no formal occupation. Bites occurring outside the African continent were always the result of individuals getting accidentally bitten while maintaining the captive snake, and were more likely to be published [21–23, 25–31, 33, 34]. This over representation of captive bite limits our understanding of puff adder envenoming in the more common rural environment. These geographic differences were also evident from the most commonly bitten body part, which was the lower limb in 66% (23/35) of all puff adder bites that happened on the African continent (case series and literature review), and the upper limb in 82% (14/17) of puff adder bites occurring outside the African continent (literature review only) [13–15, 18, 20–31, 33, 34].

### Biting snake identification

Despite the broad geographic coverage of puff adders, very few cases of confirmed puff adder bites have been reported in the literature. This is likely due to challenges in identifying the biting species in the field. Definitive identification requires the use of immunological laboratory assays, or a herpetologist to assess a picture, or parts of the biting snake. This infrastructure and expertise are not always present in areas where the most snakebites occur. In a recent review, snakebite identification by immunoassay or a herpetologist was only used in 39 and 23 of 150 publications on snakebite, respectively[35]. Most publications used less objective methods such as verbal description by the patient or bystanders, and clinical features of envenoming. This is particularly relevant for puff adder envenoming because studies in Ethiopia[36], Kenya[37, 38], Tanzania[39], Zimbabwe[3, 40], and South Africa[41] have reported high burdens of envenoming by puff adder, representing up to 75% of snakebites in some studies. Unfortunately, the method of biting snake identification was not consistently reported. This is evident in our case series where the biting snake species was objectively confirmed in two out of 15 cases. There still exists huge gaps in the diagnosis of snakebite envenoming and in the identification of the responsible biting species, and efforts to address these limitations would have the added benefit of reducing the time to treatment and improving patient outcomes.

### Local features

The most common local symptoms of puff adder envenoming were pain and swelling. Fang marks were a common feature after a bite, they were visible in all patients in our case series and were mentioned in 50% of studies in our review. Two recent reviews on dry bites did not find a case of a dry puff adder bite[42, 43], partly influenced by health seeking behaviours. Although the relevance of fang marks in the diagnosis of snakebite envenoming is debated, puff adders have large fangs, measuring 16-18 mm in length, and this muscular snake species can strike with force, likely increasing the chances of envenoming[1]. The occurrence of dry bites after bites by other medically important snake species, including members of the *Echis* and *Naja* genera, have been reported to range between 8 and 30%[44, 45]. Nonetheless, as with all other cases of snakebite, patients presenting with a suspected puff adder bite, with or without fang marks, should be hospitalised and kept under close observation for at least 24 hours.

Local swelling and pain are well-described symptoms, and they are likely due to the effects of snake venom metalloproteinase (SVMP), phospholipases A_2_ (PLA_2_) and other cytotoxic venom toxins[46, 47]. They cause local tissue damage by hydrolysing extracellular matrix components such as collagens, hyaluronic acid, and proteoglycans[47–49]. Most venom components are absorbed through the lymphatic system[50], and as with many other species, this likely explains the tender swollen lymph nodes proximal to the bite site. Other less well understood local symptoms include blister formation, necrosis, and erythema. The variability in manifestation between patients, and the specific mechanisms underlying these symptoms remain poorly understood.

### Systemic symptoms

In our case-series, haemorrhage was the most commonly reported systemic feature of puff adder envenoming, followed by coagulopathy and hypotension. From the review, hypotension, fever, and anaemia were the most commonly reported systemic features of puff adder envenoming. These three features – haemorrhage, coagulopathy, and hypotension, did occur independently but could also be linked. The haemorrhage from puff adder bites is said to most often be local, from the bite site, and can persist for up to 48 hours after the bite[16]. *Bitis arietans* venom also contains proteins and peptides that can impair platelet function [51–59] or directly damage platelets, resulting in thrombocytopenia [60–62]. A combination of platelet inactivation and depletion therefore likely contribute to the prolonged local haemorrhage observed in puff adder envenoming.

We also described cases of less clinically obvious haemorrhage in our review. Two patients had subcutaneous haemorrhage into the arm and lower back after a bite to the finger[33, 34], and one patient developed a haematoma in the jaw after a bite to the foot[19]. These accounts are suggestive of more than perturbed platelet function. SVMPs and snake venom serine proteases (SVSPs), both of which are found in puff adder venom, can hydrolyse type I and IV collagen in the basement membranes of blood vessels (mostly capillaries), causing microvascular damage, increasing vascular permeability and leading to the extravasation of blood into subcutaneous or other concealed spaces[47, 48, 63]. An autopsy in one of the fatal cases of puff adder envenoming revealed haemorrhages in the walls of the small intestine and into the adventitia of the aorta[4]. The contribution of coagulopathy in these cases of concealed haemorrhage is uncertain, and a future robust assessment of coagulation parameters is needed for further clarification. *Bitis* spp. venom also contains SVSPs that act as thrombin-like enzymes and therefore also possess fibrinogenolytic activity and can cause coagulopathy[46, 51, 64]. This coagulopathy is less severe when compared to the venom induced consumptive coagulopathy (VICC) observed after envenoming by the related saw-scaled vipers (*Echis* spp.). The highest serially measured INR in our scoping review was 2.93[33], much lower than the unrecordable INRs (i.e. >7) frequently seen with *Echis* spp. envenoming[65]. We did not find any evidence of complete fibrinogen consumption, as when measured, fibrinogen levels were normal in the two studies that reported serial tests, suggesting a more prominent role of vessel wall degradation and platelet dysfunction, than consumption of coagulation factors and ensuing coagulopathy. However, these assays were performed in a very limited number of patients and results should be interpreted with caution.

Hypotension in patients with puff adder envenoming can be a direct action of venom toxins, or an indirect consequence of haemorrhage and limb swelling. *Bitis* spp. venom contains large amounts of adenosine[66] and also bradykinin potentiating peptides[67] and kinin-releasing SVSPs[68] that cause the release of bradykinin and other vasoactive mediators which inhibit angiotensin converting enzyme and lead to haemodynamic disturbance[64, 66]. There is also evidence that SVMPs in puff adder venoms can directly generate the vasodilator angiotensin 1-7[69]. Another known component of puff adder venom, vascular endothelial growth factor[46], can also be a cause of hypotension. Hypotension can also result from the reduction in circulating blood volume following microvascular damage and extravasation of blood from the capillaries, or extravasation of fluid into swollen limbs or body parts. The acute nature of hypotension from puff adder envenoming potentially carries a high risk of fatality from shock[4], making it an important feature of envenoming. This should be actively sought out and managed, because a timely fluid bolus and appropriate antivenom can quickly reverse hypotension in patients with puff adder envenoming and improve the prognosis[4, 25, 27, 34]. Some authors recommend serial and close monitoring of the blood pressure and pulse in patients with puff adder envenoming for 48 hours[4].

Fever was another common systemic feature in our review, often accompanied by local signs of inflammation such as swelling and pain but also occurring without signs of wound infection or necrosis. *Bitis arietans* venom has been shown in-vivo to strongly activate the inflammatory process, with the induction and production of inflammatory mediators – cytokines, chemokines and eicosanoids[68, 70]. This marked production of potent endogenous pyrogens such as IL-1β, IL-6 and TNF-α, can interact directly with the anterior hypothalamus, through a hierarchy of neural structures, and raise the temperature setpoint, causing fever[71, 72]. They also activate the endothelium, inducing vasodilation and increased vascular permeability, and activate hepatocytes, inducing the production of acute phase proteins which amplify the inflammatory process[68, 73].

While a discussion of treatment options and outcomes are beyond the scope of this paper, we would like to highlight the use of fasciotomies in four out of six patients with clinical suspicion of compartment syndrome reported in the literature. The role of surgery in the management of puff adder envenoming is uncertain and compartment syndrome is rare. The extreme swelling after puff adder envenoming means that clinical signs such as pulselessness, pain, paraesthesia, and cold extremities are common and can cause overdiagnosis of compartment syndrome; these should always be accompanied by measurements of intra-compartmental pressure, or by ultrasound imaging to rule out vascular compromise. The two patients who were managed conservatively made a full recovery, suggesting timely administration of antivenom, alongside supportive treatment such as intravenous fluids may be sufficient.

Limitations in the scoping review methodology, especially our limited search of grey literature sources, may have contributed to the under reporting of puff adder bites in African countries, and reduced the validity of the features of envenoming reported in this study. Publication and observer bias may also skew our understanding of the pathophysiology of puff adder envenoming towards envenoming by captive snakes over wild snakes. Limitations in the accurate identification of the biting snake and in serial laboratory assays hinder progress in forming a global understanding of the pathophysiology of puff adder envenoming. Nonetheless, by combining data on 52 cases of puff adder envenoming we enabled the synthesis of common local and systemic signs, revealed key features of envenoming that require different therapeutic approaches, and highlighted some differences between envenoming by captive and wild puff adders.

## Conclusion

The puff adder is a medically important snake in Africa and the middle East, yet there are still gaps in the understanding of envenoming after a bite. We demonstrate that the most common local symptoms of puff adder envenoming are pain and swelling, while fang marks are typically easily visible after a bite. Systemic features include haemorrhage, coagulopathy, and hypotension, which may have a shared pathophysiology, and require timely recognition and correction to avoid fatal outcomes. Fever is a less commonly reported but frequent feature of puff adder envenoming, and it can be an early indicator of envenoming in patients with subtle local or systemic signs. Definitive identification of the biting snake, and clinical research with serial laboratory assays, is crucial to further understanding the pathophysiology of envenoming by this medially important snake species.

## Funding statement

This study was funded by the National Institute for Health Research grant awarded to RAH, GOO, NRC and DGL entitled ‘NIHR Global Health Research Group on African Snakebite Research’ (#16.137.114). The funders had no role in study design, data collection and analysis, decision to publish, or preparation of the manuscript. The views expressed are those of the author(s) and not necessarily those of the NIHR or the Department of Health and Social Care.

## Data Availability

The data used for writing and interpreting our findings are available in the manuscript

## Acknowledgements

We would particularly like to thank the patients who participated in this study. We also thank the staff in Chemolingot and Mwingi sub county hospitals for their contributions in making this study a success. We greatly appreciate the support from the National and County Ministries of Health, especially Dr Sultani Matendechero for pushing the snakebite agenda at the highest levels of government and advocating for reducing the burden of snakebite in Kenya. We sincerely thank the other members of K-SRIC and KIPRE for their valuable contributions towards the conduct of this study.

## Notes

### Competing Interest Statement

The authors have declared no competing interest.

### Author Declarations

Ethical approval for the case series portion of this study was obtained from the Kenyatta National Hospital - University of Nairobi and Ethics Research Committee, Nairobi, Kenya (P149/03/2020) and the Liverpool School of Tropical Medicine Research Ethics Committee (Research Protocol 18-058).

